# Guided relaxation-based virtual reality versus distraction-based virtual reality or passive control for postoperative pain management in children and adolescents undergoing Nuss repair of pectus excavatum: protocol for a prospective, randomized, controlled trial (FOREVR Peds trial)

**DOI:** 10.1101/2020.09.11.20192708

**Authors:** Vanessa A. Olbrecht, Sara E. Williams, Keith T. O’Conor, Chloe O. Boehmer, Gilbert W. Marchant, Susan M. Glynn, Kristie J. Geisler, Lili Ding, Gang Yang, Christopher D. King

## Abstract

**Introduction:** Virtual reality (VR) offers an innovative method to deliver nonpharmacological pain management. Distraction-based VR (VR-D) using immersive games to redirect attention has shown short-term pain reductions in various settings. To create lasting pain reduction, VR-based strategies must go beyond distraction. Guided relaxation-based VR (VR-GR) integrates pain-relieving mind-body based guided relaxation with VR, a novel therapy delivery mechanism. The primary aim of this study is to assess the impact of daily VR-GR, VR-D, and 360 video (passive control) on pain intensity and opioid consumption. We will also assess the impact of these interventions on pain unpleasantness, anxiety, and benzodiazepine consumption. The secondary aim of this study will assess the impact of psychological factors (anxiety sensitivity, pain catastrophizing) on pain following VR.

**Methods and analysis:** This is a single center, prospective, randomized, clinical trial. Ninety children/adolescents, ages 8 to 18 years, presenting for Nuss repair of pectus excavatum will be randomized to 1 of 3 study arms (VR-GR, VR-D, 360 video). Patients will use the Starlight Xperience (Google Daydream) VR suite for 10-minutes. Patients randomized to VR-GR (n=30) will engage in guided relaxation/mindfulness with the Aurora application. Patients randomized to VR-D (n=30) will play 1 of 3 distraction-based games, and those randomized to the 360 video (n=30) will watch the Aurora application without audio instructions or sound. Primary outcomes are pain intensity and opioid consumption. Secondary outcomes include pain unpleasantness, anxiety, and benzodiazepine consumption.

**Ethics and dissemination:** This study follows SPIRIT guidelines. The protocol was approved by the Cincinnati Children’s Hospital Medical Center Institutional Review Board. The trial has not yet begun recruiting (recruitment to begin July 2020). Written informed consent will be obtained for all participants. All information acquired will be disseminated via scientific meetings and published in peer-reviewed journals.

**Trial registration number:** ClinicalTrials.gov NCT04351776, registered April 3, 2020.

**ARTICLE SUMMARY:** Strengths and limitations of this study

- This is a prospective, randomized clinical trial, which provides the best clinical evidence and support for VR as an intervention.
- This is the first study examining the use of VR-based interventions in a postoperative pediatric population.
- Due to the nature of the study, it cannot be blinded.
- One limitation is the specific patient population being studied: children and adolescents between the ages of 8 and 18 years undergoing Nuss repair of pectus excavatum. Patient selection may limit generalizability of findings.
- A second limitation is the conduction of the study at an academic, tertiary care, pediatric hospital; as such, these results may not be generalizable to patients in other clinical settings.

## INTRODUCTION

### Background and rationale

Children and adolescents with pain are at risk of opioid abuse,^1^ and many are initially exposed to narcotics prescribed to treat pain.^2^ Pectus excavatum, a depression of the anterior chest wall, is often corrected via the Nuss repair, a minimally invasive procedure in which a bar(s) is inserted beneath the sternum and flipped to elevate the chest.^3^ Although minimally invasive, this procedure is associated with significant postoperative pain.^4^ Despite efforts at multimodal therapy, the percentage of patients experiencing severe pain after surgery has not changed over the last 20 years.^5, 6^ Multimodal pain management requires the exploration of safe, effective, nonpharmacological strategies that reduce pain and opioid consumption.^7^

Virtual reality (VR) may offer a safe, innovative, nonpharmacological tool with the potential to decrease pain and medication consumption. Children and adolescents are at risk of persistent pain and opioid use after surgery.^8, 9^ While this risk is well documented in adults, few studies address this topic in children.^10^ Existing pediatric studies have identified an approximately 20% incidence of persistent postsurgical pain beyond what is expected from surgery alone.^11^ While 80% of these patients recover within one month, 20% maintain a reduced quality of life secondary to persistent pain.^11^ A recent retrospective study of opioid-naïve surgical patients found persistent opioid use in 4.8% of adolescents vs. 0.1% in a matched, nonsurgical cohort.^8^ Using opioids for as little as 5 days increases the risk of long-term use.^12^ However, the consequences of ineffective postoperative pain management are significant and associated with increased morbidity, poorer physical functioning, longer recovery, and higher economic cost.^13, 14^ As such, novel, nonpharmacological methods to treat pain can both improve analgesia after surgery and decrease opioid exposure, a risk factor for future addiction.^1, 2^

VR provides an immersive, multisensory, three-dimensional (3D) environment that enables individuals to have modified experiences of reality by creating a sense of “presence,” making it an excellent candidate for distraction-based therapy.^15^ Distraction-based virtual reality (VR-D) has been used during painful procedures, the postoperative period, and labor to help decrease pain by redirecting attention.^16-28^ These studies show short-term decreases in pain, but this transient reduction is insufficient to treat prolonged acute pain experiences,^29, 30^ including postoperative pain. Comparatively, nonpharmacological alternatives that utilize mind-body based therapies delivered in a traditional format, like relaxation and slow breathing, are able to decrease anxiety and pain in children undergoing surgery.^31^ However, despite their efficacy, these therapies are fraught with challenges, such as barriers to accessing care, high cost, need for multiple visits, and provider shortages.^32^ VR can increase accessibility to these mind-body therapies and enhance acceptability, motivation, and adherence in pediatric patients compared to methods without VR.^33^ Combining strategies of traditional mind-body therapies, like relaxation and slow breathing, with the immersive nature of VR opens new possibilities for multimodal analgesia in the pediatric population and has the potential to simultaneously minimize acute postoperative pain and opioid consumption. Guided relaxation-based VR (VR-GR) is a promising mechanism to deliver mind-body based therapy, improve postoperative pain control, and avoid challenges common with mind-body therapies delivered in the traditional format.

We have designed a prospective, randomized, clinical trial to assess the efficacy of VR-GR to decrease pain, anxiety, and opioid consumption in children and adolescents undergoing Nuss repair of pectus excavatum and hypothesize that VR-GR will be more effective at reducing pain, anxiety, and opioid consumption in this population than VR-D or a passive control.

### Objectives

The primary objective of this study is to determine the impact of VR-GR on pain intensity and opioid consumption in children and adolescents undergoing Nuss repair of pectus excavatum compared to VR-D and 360 video both during the hospitalization and up to one month following discharge. We will also assess the impact of VR-GR on pain unpleasantness, anxiety, and benzodiazepine consumption compared to VR-D and 360 video. The secondary objective of this study is to determine the role of anxiety sensitivity and pain catastrophizing on changes in pain and anxiety following VR-GR, VR-D, and 360 video both during hospitalization and 1-month post discharge in this same patient population using standard questionnaires.

## METHODS AND ANALYSIS

The FOREVR Peds study is a single center, prospective, unblinded, randomized clinical trial with three groups: a daily, 10-minute session of VR-GR, VR-D or 360 video in children and adolescents between the age of 8 and 18 years undergoing Nuss repair of pectus excavatum. The primary objective is to determine the impact of VR-GR on pain intensity and opioid consumption compared to VR-D and 360 video. Patient recruitment has not yet begun, and we anticipate a total study duration of two years. We anticipate patient recruitment to begin in July 2020. This study protocol complies with the SPIRIT Statement as well as the CONSORT Statement (Figure 1). The study was registered at ClinicalTrials.gov (NCT04351776) on April 3, 2020 and all trial registration data can be found on the ClinicalTrials.gov website.

**Figure 1.**
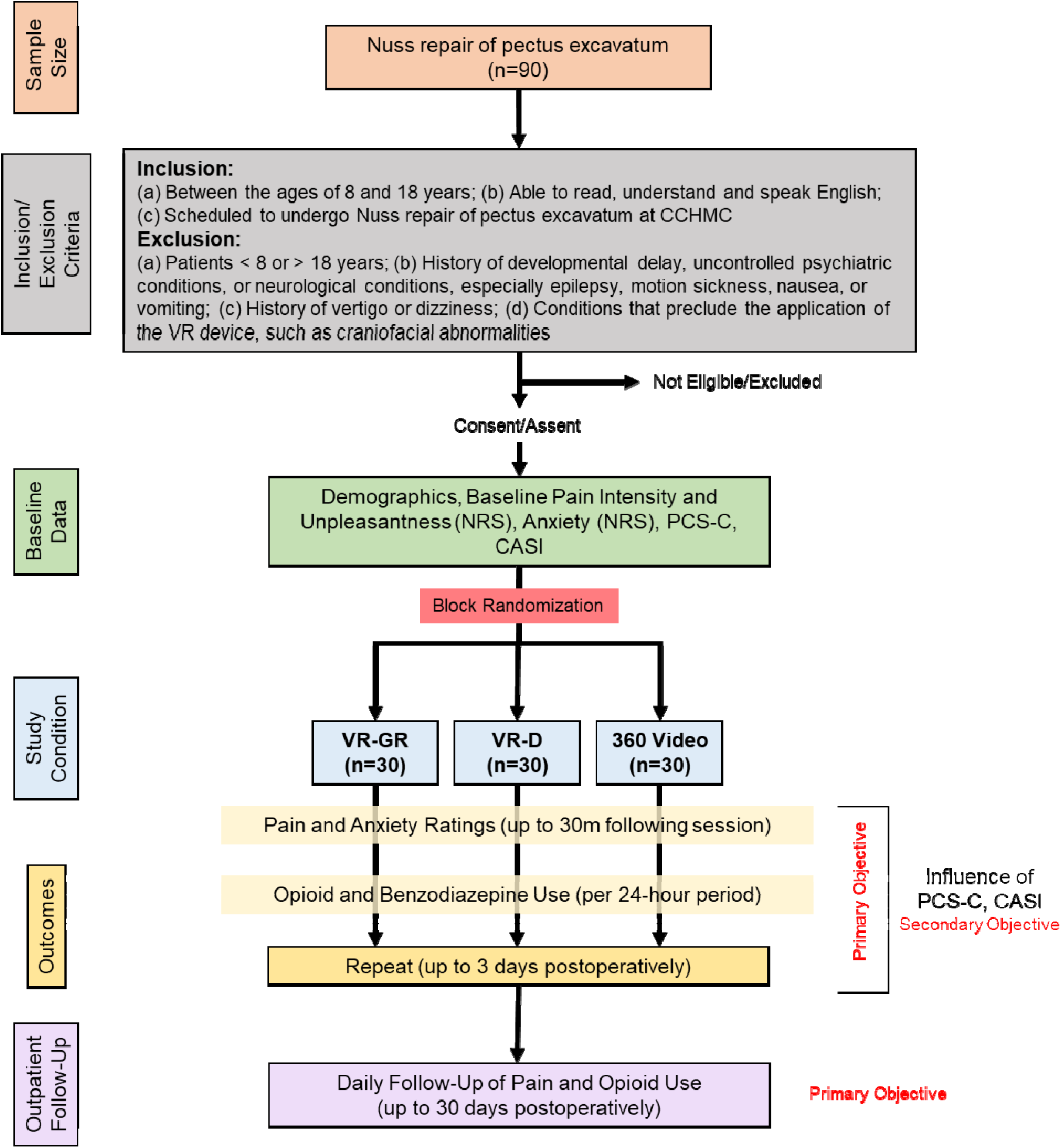
Study flow chart (CONSORT Diagram)

### Study setting

Cincinnati Children’s Hospital Medical Center (CCHMC), a tertiary care, academic, pediatric hospital.

### Study design

This is a single-center, prospective, randomized clinical trial of children and adolescents with acute postoperative pain following Nuss repair of pectus excavatum to assess the impact of multiple VR-GR sessions on pain and medication utilization in relation to patient anxiety and pain catastrophizing. Figure 1 summarizes the study design. We will assess the acute and longterm impact of each intervention on changes in pain intensity, pain unpleasantness, anxiety, and opioid and benzodiazepine consumption during hospitalization and following discharge. Figure 2 summarizes this experimental design. All patients are managed postoperatively via the pectus surgery pain management protocol, which standardizes all non-controlled medications received by patients. Patients enrolled in this study will be managed per this protocol (standard care) and will receive the additional intervention of VR or 360 video.

**Figure 2.**
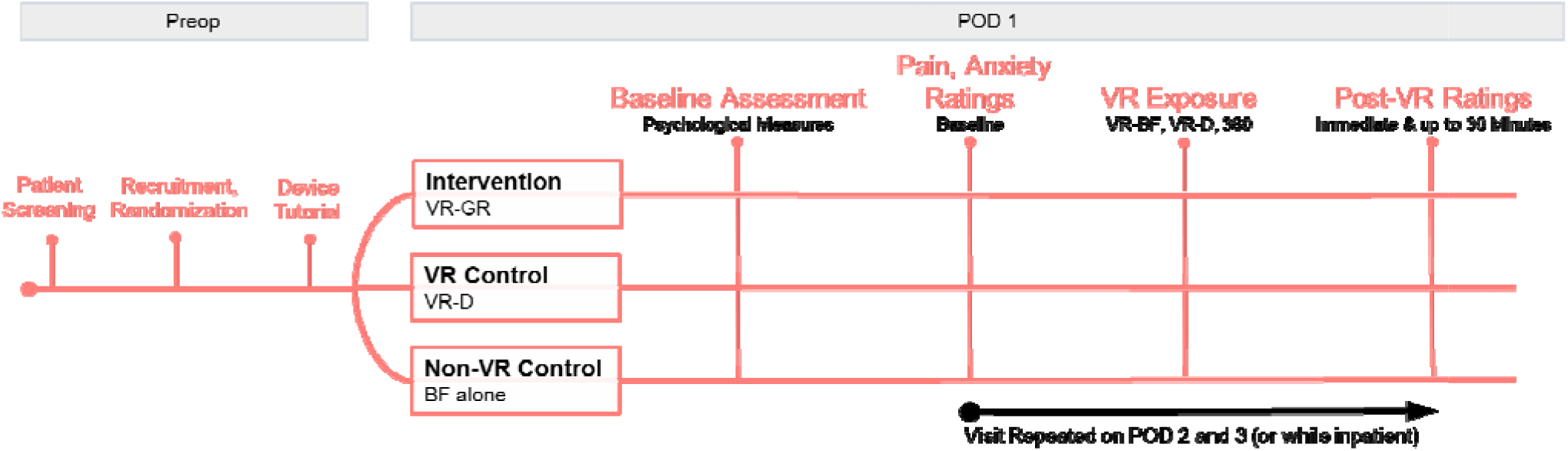
Experimental design of the study.

### Outcome measures

#### Primary outcomes

Our primary outcome is pain intensity and opioid consumption following daily VR-GR, VR-D, and 360 video in our population during hospitalization and up to 1-month post-discharge.

#### Secondary outcomes

Our secondary outcomes are pain unpleasantness, anxiety, and benzodiazepine consumption following daily VR-GR, VR-D, and 360 video in our population during hospitalization and up to 1-month following discharge. We will also assess the impact of pain catastrophizing and anxiety sensitivity on these outcomes.

### Participants

We will recruit 90 adolescents (30 per group) between the age of 8 and 18 years undergoing Nuss repair of pectus excavatum. Eligibility criteria have been chosen to correspond with our prior work and result in a population with whom our group has substantial experience.

#### Inclusion Criteria

Patients will be included based on the following criteria: (a) Between the ages of 8 and 18 years; (b) Able to read, understand, and speak English; (c) Scheduled to undergo Nuss repair of pectus excavatum at CCHMC.

#### Exclusion criteria

Patients will be excluded for the following reasons: (a) Patients < 8 or > 18 years of age at the time study enrollment; (b) History of significant developmental delay, uncontrolled psychiatric conditions, or significant neurological conditions, including epilepsy, severe motion sickness, or active nausea/vomiting; (c) Conditions that preclude application and use of the VR device, including craniofacial abnormalities.

### Randomization

Eligible patients will be randomly assigned in a 1:1:1 fashion to the following three study groups: VR-GR, VR-D, and 360 video (passive control). Block randomization will be done using REDCap (https://www.project-redcap.org/), a secure web application for building and maintaining secure databases and surveys. We anticipate that randomization will allow for equal distribution of demographic characteristics among the three groups. We will consider stratification by age, if necessary, in the analysis.

### Interventions

All patients will use the VR device and software from the Starlight Children’s Foundation, the Starlight Xperience device (Google Daydream). This VR device is commercially available and is not FDA regulated. The Google Daydream is an all-in-one headset, so no additional hardware is required to deliver the VR experience. A set of headphones, included with the headset, is used to deliver audio instructions and sound, creating a fully immersive experience. Patients will be visited daily to undergo a single, 10-minute session with the VR headset.

#### VR-GR (intervention)

Patients randomized to the VR-GR group will use the Aurora application to receive relaxation/mindfulness content. This application acts as an escape for patients as well as a tool to teach slow breathing and relaxation techniques. Patients are transported to an alpine meadow with dynamic daytime, and later, nighttime scenery. With the help of a 10-minute narrative, participants are guided to sync their breathing with their surroundings: the rise and fall of a floating butterfly during the day and the movement of the northern lights in the sky at night.

#### VR-D (active control)

Patients randomized to the VR-D group will choose 1 of 3 distraction-based games: Space Pups, Pebbles the Penguin, or Wonderglade. Each provides a similar distraction-based experience for the user. 1) *Space Pups:* user controls an astronaut space puppy and works to collect treats to the beat of the music. 2) *Pebbles the Penguin:* user controls a penguin sliding down a mountain and works to collect shiny pebbles to unlock new power-ups. 3) *Wonderglade:* 5 different carnival-themed mini-games like basketball, miniature golf, and racing.

#### 360 video (passive control)

Patients will view a 360 video of a nature scene like the Aurora application but will not receive a guided tutorial on how to relax and sync their breathing with the application. They will also not receive any audio instructions or sound, decreasing the fully immersive experience.

### Patient recruitment

On average, 125 to 150 Nuss repairs are performed at CCHMC annually. We plan to enroll a total of 90 patients. Patients scheduled to undergo Nuss repair of pectus excavatum will be recruited continuously throughout the course of the study until enrollment targets are met. We anticipate recruiting two patients per week given our surgical volume. We will receive notification of all Nuss repair surgery bookings by the surgery schedulers to identify possible participants, allowing for eligible patients to be identified greater than 1 week prior to surgery. The operating room schedule as well as the surgical patient list will be screened for eligible patients based upon age criteria. Patients meeting age criteria will undergo screening of their available electronic medical record to assess study eligibility. Eligible patients will be approached on the day of surgery. If patients wish to participate, appropriate consent (and assent for patients ≥ 11 years of age) will be obtained and eligibility criteria will be verified. Patients will be block randomized (1:1:1 ratio using REDCap) to VR-GR (intervention), VR-D (active control), and 360 video (passive control). Patients will receive a tutorial on the VR device at the time of enrollment. Demographic, health information, and medical history will be recorded and documented in the REDCap database. Patients will be offered a small stipend for participation to help increase recruitment and adherence.

### Study visits

Patients will be visited daily to undergo a single, 10-minute session. Prior to the first session, patients will complete two validated questionnaires to assess baseline trait measures: the Pain Catastrophizing Scale for Children (PCS-C)^34^ and the Child Anxiety Sensitivity Index (CASI).^35^ They will also complete a health history questionnaire and a baseline pain intensity, pain unpleasantness, and anxiety rating will be obtained using the Numerical Rating Scale (NRS).^36, 37^ Pain intensity, pain unpleasantness, and anxiety ratings will be repeated immediately, 15 minutes, and 30 minutes following session completion. Patients typically remain in the hospital for 3 to 4 days following Nuss repair. During their inpatient stay, participants will have daily study visits, repeating the same process as the first session; patients will not repeat the PCS-C or CASI surveys. At the last visit, patients will be given a satisfaction survey to gather qualitative feedback about the VR experience.

### Data collection

For each eligible participant, data will be collected from patient history/interview and the electronic medical record in a standardized case report form in the REDCap system by a clinical research coordinator (CRC) or student who maintain CITI training in accordance with our local institutional review board (IRB) under the direct supervision of the principal investigator (PI). Total opioid and benzodiazepine usage will be collected from the electronic medical record for 24 hours after each session. All medication consumption will be collected for assessment of non-opioid analgesics and to ensure consistency with the pectus pain management protocol. To assess pain intensity and unpleasantness after hospital discharge, patients will use a daily log to record pain scores using the NRS for one month. We will use eCAP (electronic capture pill dispenser, https://www.informationmediary.com/nfc-smart-packaging-devices/ecap-smart-pill-bottle/) to document medication consumption. Weekly reminders will be sent using Twilio, and telephone follow-up will be done at two weeks and one month to help improve patient adherence.

Prescription cross-verification will be done using controlled substance reporting databases for Ohio, Kentucky, and Indiana (OARRS, KASPER, and INSPECT, respectively) to verify data collected from patient logs and eCAP.

### Measurements

a) Pain intensity, pain unpleasantness, and anxiety ratings will be assessed using the NRS.^36, 37^ b) Pain catastrophizing will be assessed using PCS-C.^34^ c) Anxiety sensitivity will be assessed using CASI.^35^ d) Total opioid and benzodiazepine usage will be collected from EPIC for 24 hours after each session and up to 1-month post-hospital discharge via eCAP. All medication consumption will be collected for assessment of non-opioid analgesics and converted to milligram per kilogram per day. Table 1 summarizes the measurements used in the study.

**Table 1.** Scales and questionnaires used in the study.

| Table 1. Scales and questionnaires for the study |
| --- |
| **<u>Numerical Rating Scale (NRS).</u>** Numerical rating scale where children are asked to give a number on a scale of 0 to 10 of how bad their pain hurts, with 0 being no pain and 10 being the worst pain of their life. |
| **<u>Pain Catastrophizing Scale for Children (PCS-C).</u>** Children rate 13 items assessing rumination, magnification, and helplessness related to thoughts about pain. PCS summary scores can be interpreted as low (0 to 14), moderate (15 to 25), and high ($\geq 26$). Internal reliability for our VR-D pilot data was 0.94 (Cronbach's $\alpha$). |
| **<u>Child Anxiety Sensitivity Index (CASI).</u>** 18-item self-report tool designed to measure symptoms of anxiety in children and adolescents, with total scores ranging from 18-54. Internal reliability for our VR-D pilot data was 0.84 (Cronbach's $\alpha$). |

### Sample size calculation

Sample size calculation is based on preliminary data assessing the impact of VR-D to affect changes in pain intensity in children and adolescents following surgery (unpublished). Preliminary data showed that the average change in pain intensity across time was -1, with standard deviation (SD) 1.2 and correlation between measurement pairs of 0.88. Assuming similar results in the passive control group, sample size of 30/group will have 80% power to detect differences in mean changes of one between VR-GR and the two control groups. We expect a difference of ≥1 between VR-GR and VR-D to emerge with multiple sessions as proposed with this study. Significance (alpha) is 0.025 to control for two comparisons. We will recruit 90 patients, 30 per group.

### Statistical analysis

Statistical analysis will be done with SAS 9.4 (Cary, NC). Descriptive statistics will be calculated and summarized (*<u>continuous</u>*: mean ± SD; *<u>categorical</u>*: frequency %). Prior to analysis, assumption of normality will be assessed for continuous variables and corrected using log transformation when appropriate. Bonferroni correction will be made as appropriate for comparisons. Change from baseline for primary and secondary outcomes will be tested for normality and deviation from zero using paired tests (t-test or signed-rank, as appropriate) at individual time points after interventions. Change from baseline will be compared between groups using two-sample t-test or Wilcoxon rank-sum test (between two groups, i.e., VR-GR vs. VR-D and VR-GR vs. 360 video) and ANOVA or Kruskal-Wallis test (across three groups) at individual time points after the sessions. Mixed effects models for repeated measures with baseline value, intervention group, time (0, 15, 30 minutes after intervention), and group and time interaction will be used to test the hypothesis that VR-GR reduces pain, anxiety, and opioid and benzodiazepine consumption more than controls. Pain and opioid use 1-month post-discharge will be compared between intervention groups using ANOVA (with adjustment of possible covariates) or Kruskal-Wallis test, as appropriate, based on data distribution.

Anxiety sensitivity (or pain catastrophizing) will be dichotomized using the sample median (or tertiles depending on distribution) and its effect on response to intervention (change in pain intensity from baseline) will be tested using the two-sample t-test or Wilcoxon rank-sum test, as appropriate at individual time points (0, 15, 30 minutes) after intervention. Mixed effects models for repeated measures (change in pain intensity from baseline) with high or low anxiety sensitivity (or pain catastrophizing) group, time (0, 15, 30 minutes after intervention), and group and time interaction will be used to test the hypothesis that patients with greater anxiety sensitivity and pain catastrophizing will have a larger reduction in pain vs. patients with less anxiety sensitivity and pain catastrophizing. Assuming the same SD and correlation between pain intensity measurement pairs from the primary power analysis, sample size of 15/group (high vs. low anxiety or pain catastrophising dichotomized at median for the VR-GR group) will have 80% power to detect differences in mean changes of pain intensity of 1.3 between the two groups, with alpha=0.05. The same analysis will be repeated for pain unpleasantness and anxiety.

We will make every effort to ensure that at least one daily VR session will be completed for each study participant and that all data extraction will be complete to avoid missing data. We will assess missing data for all study variables. Chart review for missing data on demographics, medical history, etc. will be performed when feasible. Missing outcome data will be statistically imputed using last observation carried forward (LOCF) or multiple imputation, and a sensitivity analysis will be conducted with different imputation methods as well as without imputation.

## ETHICS SAFETY AND DISSEMINATION

### Ethics

This study is being conducted in accordance with the rules and regulations applicable to the conduct of ethical research and this study protocol has been approved by the IRB at CCHMC (IRB #2019-1090). This protocol includes clear delineation of the protocol version identifier and date on each protocol amendment submitted to the IRB; clear delineation of plans for data entry, coding, security, and storage; clear delineation of mechanisms to ensure patient confidentiality, including how personal information will be collected, shared, and maintained in order to protect confidentiality before, during, and after the trial; statements regarding who has access to data collected during this study; and a model consent form and other related documentation given to participants and/or guardians. We do not anticipate any major protocol modifications during the duration of this study.

### Safety

It is anticipated that the risk to participants in this study is minimal. The specific VR device used in this study is a minimal risk device, and because it is considered a relaxation device by the FDA, it is not regulated as a clinical device. Risks specific to VR are minimal, with the greatest risk being motion sickness and/or nausea while the headset is in place.^38^ There is a theoretical risk of inducing seizures (0.025% in a pediatric data set supplied by a similar Samsung device). We will minimize these risks by excluding patients with a history of significant neurological disorders, including epilepsy and severe motion sickness/nausea. Patients will also be explicitly instructed to remove the headset should any side effects or discomfort occur. The PI will continually monitor all risks to the participants. Weekly lab meetings will be used to address quality assurance and safety concerns with the study. Research personnel are instructed to inform the PI immediately with any safety concerns or adverse events (AEs). The IRB will also be updated when any serious AEs (SAEs) occur or when mild or moderate AEs determined to be a result from study participation occur. SAEs that are unanticipated, serious, and possibly related to study participation will be reported to the data safety monitoring committee (DSMC), IRB, and any other necessary study regulatory committee. We do not anticipate any SAEs that would require stopping this trial early. Therefore, we do not plan to conduct interim analysis for safety. This consideration will change if SAEs are reported during the study.

Although the risk to patients from this clinical trial is low, a DSMC will be utilized to monitor safety. The DSMC will be composed of three experts (clinical research, pain management, and digital technology) who are independent of the protocol. The DSMC will report to the IRB. This protocol is approved by the IRB at CCHMC in compliance with existing regulations and policies for the conduct of clinical research.

### Dissemination

Unique data will be obtained from this research and will be widely disseminated through conference presentations at national and international meetings and through publication of manuscripts in peer-reviewed publications. Participants may receive trial results if interested. All authors are eligible to participate in dissemination and we do not plan to use professional writers to disseminate study results.

### Patient and public involvement

No patients or members of the public were involved in the design, recruitment, or conduct of this study. Consideration of the burden of the intervention and time required to participate in this research was assessed during pilot data collection using VR in the acute postoperative pain population at our institution and information gathered from this pilot study helped guide the development of this clinical trial. Participants may receive information about study results if they wish via a letter describing results to participants. We will share access to the full protocol to requesting individuals/institutions.

## CONCLUSION

In summary, this is the first study to assess the efficacy of VR-GR compared to VR-D and a passive control. If this study yields beneficial results, we hope to design a multi-center, prospective, randomized clinical trial and incorporate VR-GR into multimodal analgesia in children and adolescents after surgery. Ultimately, this technology has the potential to impact care by providing remote delivery of this effective therapy and decreasing pain and opioid consumption in a variety of patient populations with pain.

## Data Availability

Data will be available upon request.

## AUTHOR CONTRIBUTIONS

VO, SW, and CK contributed to the conception of this idea, the design of the research protocol and study, and the writing of this manuscript. KO, CB, GM, SG, and KJ provided input regarding the design and implementation of the study protocol and procedures. LD and GY contributed to the design of the research protocol and the statistical analysis plan development. All authors revised and modified this manuscript. They will all approve the final version.

## DATA STATEMENT

Technical appendix, statistical code, and/or the study dataset will be available to the public following study completion.

## ACKNOWLEDGEMENTS

The authors have no acknowledgements to disclose.

